# Sleep Duration and the Risk of Metabolic Syndrome: a Systematic Review and Meta-analysis

**DOI:** 10.1101/2020.08.30.20184747

**Authors:** Jianian Hua, Hezi Jiang, Qi Fang

## Abstract

**Objective:** Epidemiologic studies have reported inconsistent findings about the association between sleep duration and metabolic syndrome. We aimed to clarify this association by method of metaanalysis.

**Methods:** Medline, Embase, CINAHL and PsycINFO databases were searched from inception to May 2020. We collected data from 235,895 participants from 9 prospective cohort studies, and 340,492 participants from 26 cross-sectional studies. Risk ratios (RR) or odds ratios (OR) were calculated separately for cohort studies and cross-sectional studies, through meta-analysis of adjusted data from individual studies.

**Results:** For cohort studies, short sleep duration was associated with an increased risk of metabolic syndrome (RR, 1.15; 95% CI, 1.05-1.25). Long sleep duration was not associated with new onset metabolic syndrome (RR, 1.02, 0.85-1.18). For cross-sectional studies, both short (OR, 1.12, 95% CI, 1.08-1.18) and long (OR, 1.06, 1.01-1.11) sleep duration was associated with high prevalence of metabolic syndrome.

**Conclusions:** Only short sleep duration was associated with an increased risk of metabolic syndrome. Future studies should address whether the association is casual and modifiable.

## 1 Introduction

Metabolic syndrome is a cluster of disorders that occur together, and it is defined as a condition consisting of at least three of the following: hypertension, high blood sugar, excessive waist circumference, higher triglyceride (TG) levels or low high-density cholesterol (HDL) levels. Metabolic syndrome is associated with adverse cardiovascular events, even after adjusted for diabetes and obesity^1,2^. Meanwhile, the prevalence of metabolic syndrome in America is up to 33%^3^. Recently, growing concern has been paid to the modifiable risk factors of metabolic syndrome^4^.

Abnormal sleep duration is known to increase the risk of a serious health outcomes, including mortality, diabetes and cardiovascular disease^5,6^. Therefore, both sleep and metabolic syndrome increase the risk of cardiovascular event. However, whether abnormal sleep duration increase the risk of metabolic syndrome remained controversial. Several meta-analyses have reported the association between sleep duration and metabolic syndrome^7-9^. However, none of the studies assessed the association in a prospective cohort design, which has less substantial bias could provide stronger support for causality^6,10,11^.

Therefore, we conducted a systemic review and meta-analysis to: (1) examine all prospective cohort studies that have assessed the effect of sleep duration on the future risk of metabolic syndrome; (2) conduct an updated meta-analysis to examine the cross-sectional association.

## 2 Materials and Methods

We performed this study according to Preferred Reporting Items for Systematic Reviews and MetaAnalysis (PRISMA) guidelines (Table S1).

Two independent researchers (JNH and HZJ) separately assessed the eligibility, extracted data, and checked the quality of the included studies. Any disagreement in screening the articles were resolved through discussion between these two, with adjudication by a third research (QF) if disagreements persisted.

### 2.1 Search strategy

A systematic search was conducted in Medline, Embase, CINAHL and PsycINFO from inception to May 2020. We used the following search terms: (1) “sleep hour” OR “sleeping hour” OR “hours of sleep” OR “sleep duration” OR “sleep time” OR “sleep length” OR “sleep period” OR “sleeping time” OR “sleep span”; (2)“metabolic syndrome” or “MetS” or “MS” or “syndrome X” or “cardiometabolic risk factor” or “insulin resistance syndrome”. Algorithms combining the two sets of search terms were applied.

### 2.2 Study selection

Studies that examined the relationship between sleep duration and metabolic syndrome were screened for eligibility, without language restriction. The inclusion criteria were: (1) the research was a cohort or a cross-sectional study; (2) the research compared individuals with “unnormal” and “normal” sleep duration on the outcome of metabolic syndrome. If one or more articles reported data from one study, the article with the largest sample size and the best design was included.

We analyzed cohort studies and cross-sectional studies separately in all analysis due to the following reasons. First, cross-sectional study could not establish a causal relationship, and it only reflected the association between sleep duration and the prevalence of metabolic syndrome^11^. Second, the non-exposed prevalence of metabolic syndrome was higher than 20%^12^, odds ratios (OR) could not be regarded as risk ratio (RR)^13^.

The inclusion of studies was conducted in two stages: (1) screening of the title and abstract and (2) screening of the full text.

### 2.3 Data extraction

The following characteristics were extracted from each eligible study: author name and publication year, study type, study location (country and continent), sample size, participants’ characteristics (age range, mean age and sex composition), exposure and outcome assessment (sleep assessment, metabolic syndrome measures and definition of long or short sleep duration) and main results.

Since definition of sleep duration varies among studies^14^, the three categories of sleep duration (short, long and normal) was extracted by one of the following two ways. For some papers, sleep duration was already divided into three categories based on cultures and ethnicities. For others, in which sleep duration was divided into more than three groups, short or long sleep duration was defined as the shortest or longest group reported in the article^6^. With regard to the main results, the adjusted estimates that reflected the most comprehensive control were extracted.

Data were extracted by two investigators (JNH and HZJ) independently. Disagreement in screening the articles was resolved by discussion between the two investigators. Consultation from a third investigator (QF) was acquired if necessary.

### 2.4 Exposure and outcome measures

With regard to the measurements of nocturnal sleep duration, two studies used objective measurement, while others used interview or questionnaire.

The diagnostic criteria of metabolic syndrome (MetS) varied between studies. 10 studies used the Third Report of the National Cholesterol Education Program’s Adult Treatment Panel III (NECP ATP-III); 4 studies used modified NECP ATP-III; 14 studies used the American Heart Association/National Heart Lung and Blood Institute (AHA-NHLBI); 3 studies used the International Diabetes Federation (IDF)^8^; 5 studies used other criteria.

### 2.5 Quality appraisal

The quality of all studies was evaluated by Newcastle-Ottawa Quality Assessment Scale (NOS)^15^. The total score ranged from 0-9 scores. For outcome category and comparability category, all the studies had similar quality. The difference between studies lied in the study design category (Table S3).

### 2.6 Data analysis

We conducted all the following analysis separately for cohort studies and cross-sectional studies, and for study-specific short and long sleep duration, respectively.

In analysis of cohort studies, hazard ratios (HRs) were regarded as risk ratios (RRs). For studies which provided only odds ratios (ORs), we calculated RRs by using the ORs and control event rates (CERs) in normal sleepers. Using random-effect models, we estimated the pooled RR and 95% CI. With regard to cross-sectional studies, we calculated the pooled OR and 95%CI using random-effect models.

Heterogeneity between the studies was assessed using Cochran Q statistics (P<0.1 indicates statistically significant heterogeneity) and *I*^2^ statistic (*I*^2^>50% indicates statistically significant heterogeneity)^16^. To examine publication bias, we used a funnel plot, the Egger regression test, and the Begg and Mazumdar test^17,18^. “Trim and fill” method was used to adjust the funnel plot, and recalculated the results^19,20^. Sensitivity analysis was performed by excluding one study at a time to test the robustness of the pooled estimates.

Subgroup analysis was conducted to explore the potential heterogeneity among cross-sectional studies. We used z test to compare summary estimates of each subgroups^21^. Univariate and multivariate meta-regression were used to study the effect of possible influential confounders, such as definition of sleep duration and study quality. For cohort studies, subgroup analysis and metaregression was not performed considering the small number of datasets included in the meta-analysis. All statistical analysis was performed by R-3.4.3^20^ and Stata 15.1 (Stata Corp, College Station, TX).

## 3 Results

### 3.1 Search results

The initial electronic search yielded 1140 articles, among which 789 were reviewed by title and abstract. 127 full-texts were retrieved. 35 studies were included in the final analysis (Figure 1).

**Figure 1.**
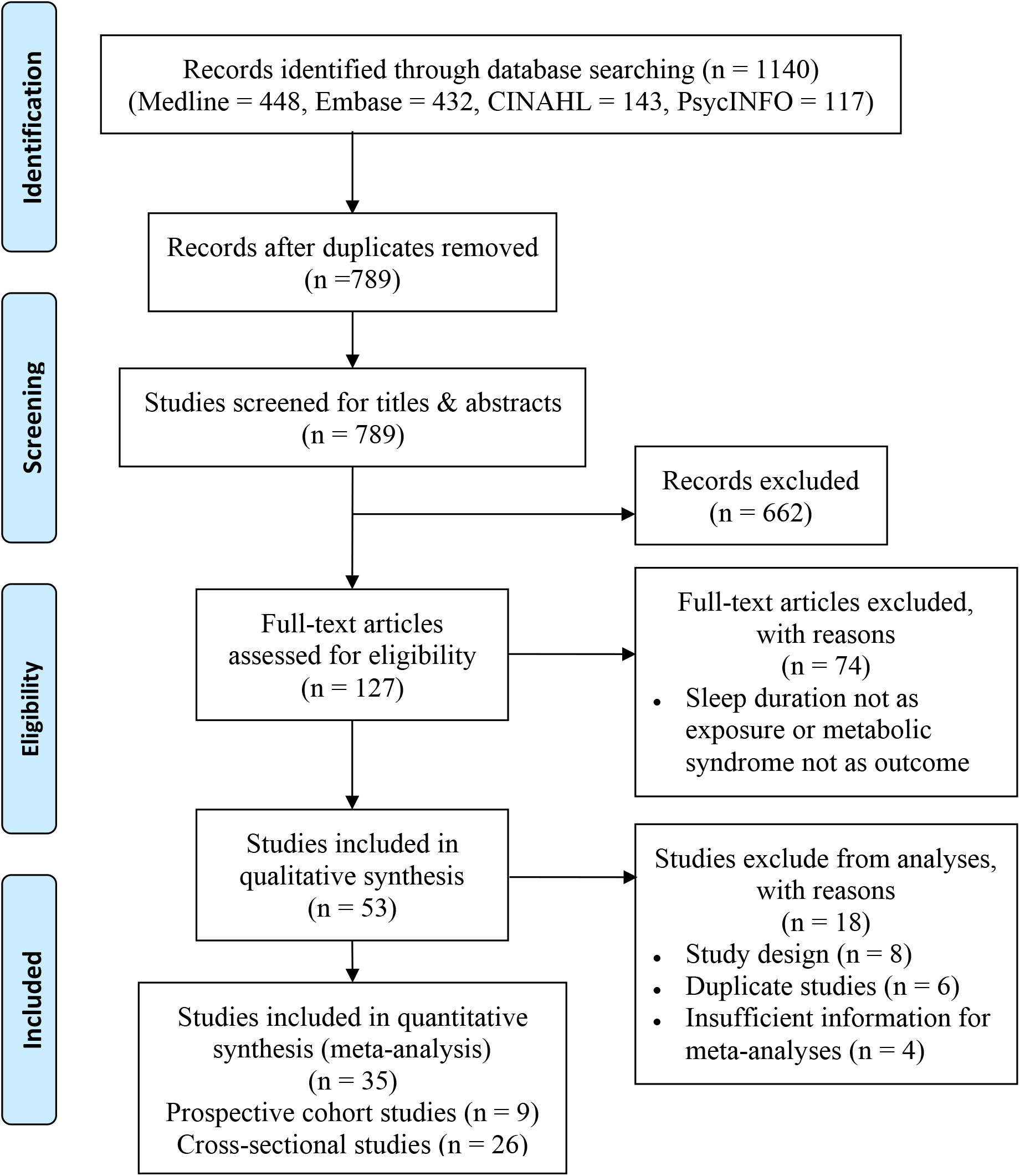
Flowchart for the included studies.

### 3.2 Characteristics of study samples

We identified nine prospective cohort studies that examined the association between sleep duration and the incident risk of metabolic syndrome, in a total of 235,895 participants. The sample size ranged from 293 to 162,121. The mean follow-up duration ranged from 2 to 8 years (Table 1).

**Table 1.**
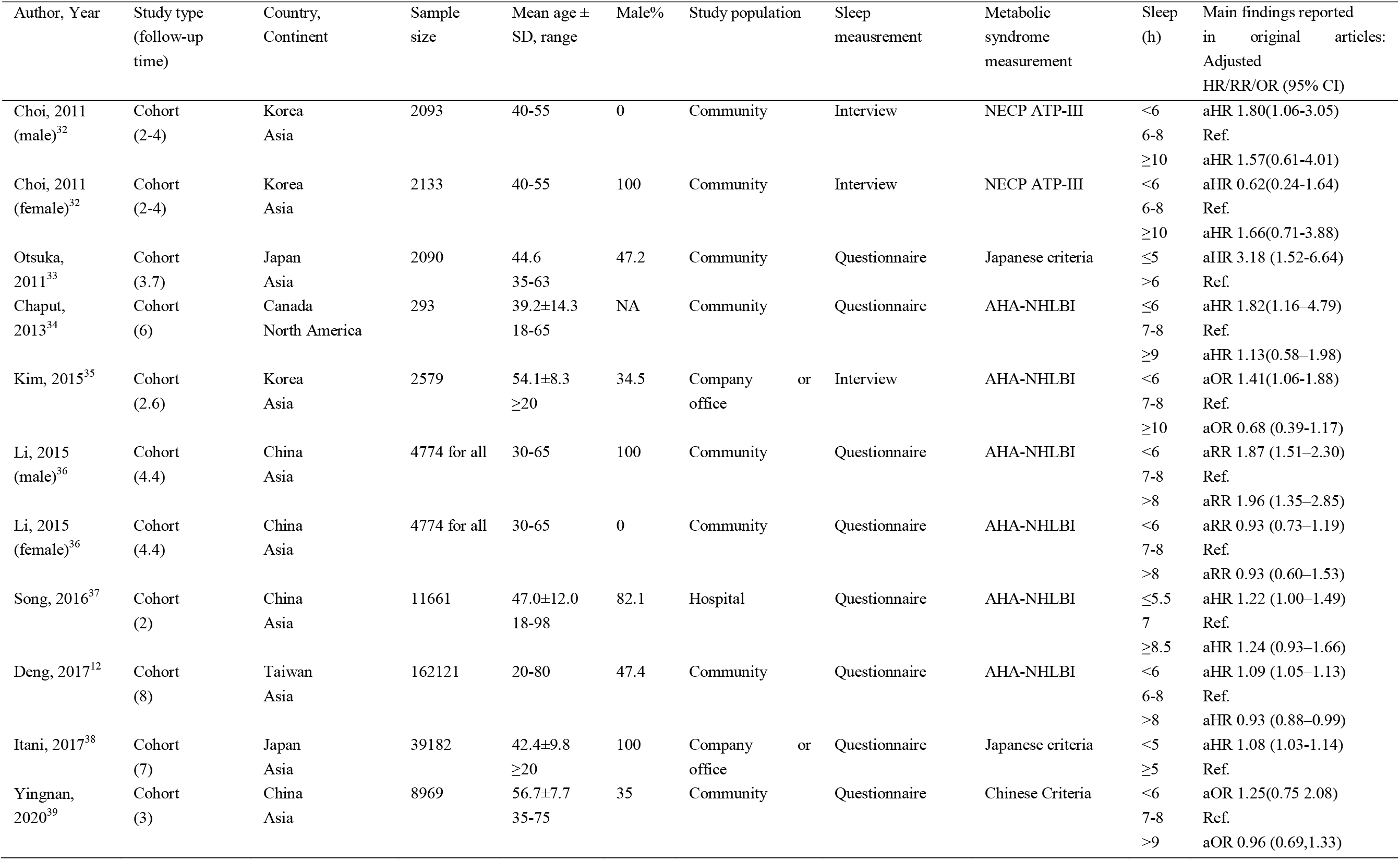
Characteristics of cohort studies.

Another twenty-six studies were cross-sectional studies, included a total of 340,492 individuals. The sample size ranged from 263 to 88,678 (Table S2).

Table 1 and Table S2 presented the characteristics of the all 35 studies. The individuals all aged above 18 year. The mean (SD) age of the individuals ranged from 31 (8.7) to 67.6 (7.3) years. The studies were conducted in five continents, while many of them were from Asia. The definitions of short or long sleep duration varied between studies. About 75% studies defined “short sleep duration” as 6 or 7 h, and about 80% studies defined “long sleep duration” as 8 or 9 h.

### 3.3 Primary analysis

#### 3.3.1 Sleep duration and the risk of metabolic syndrome

Compared with moderate sleep duration, short sleep duration was associated with a statistically significant increase in new onset metabolic syndrome, with an RR of 1.15 (95% CI=1.05-1.25, P<0.001, I^2^=63.6%, N of datasets=11; Figure 2A).

**Figure 2.**
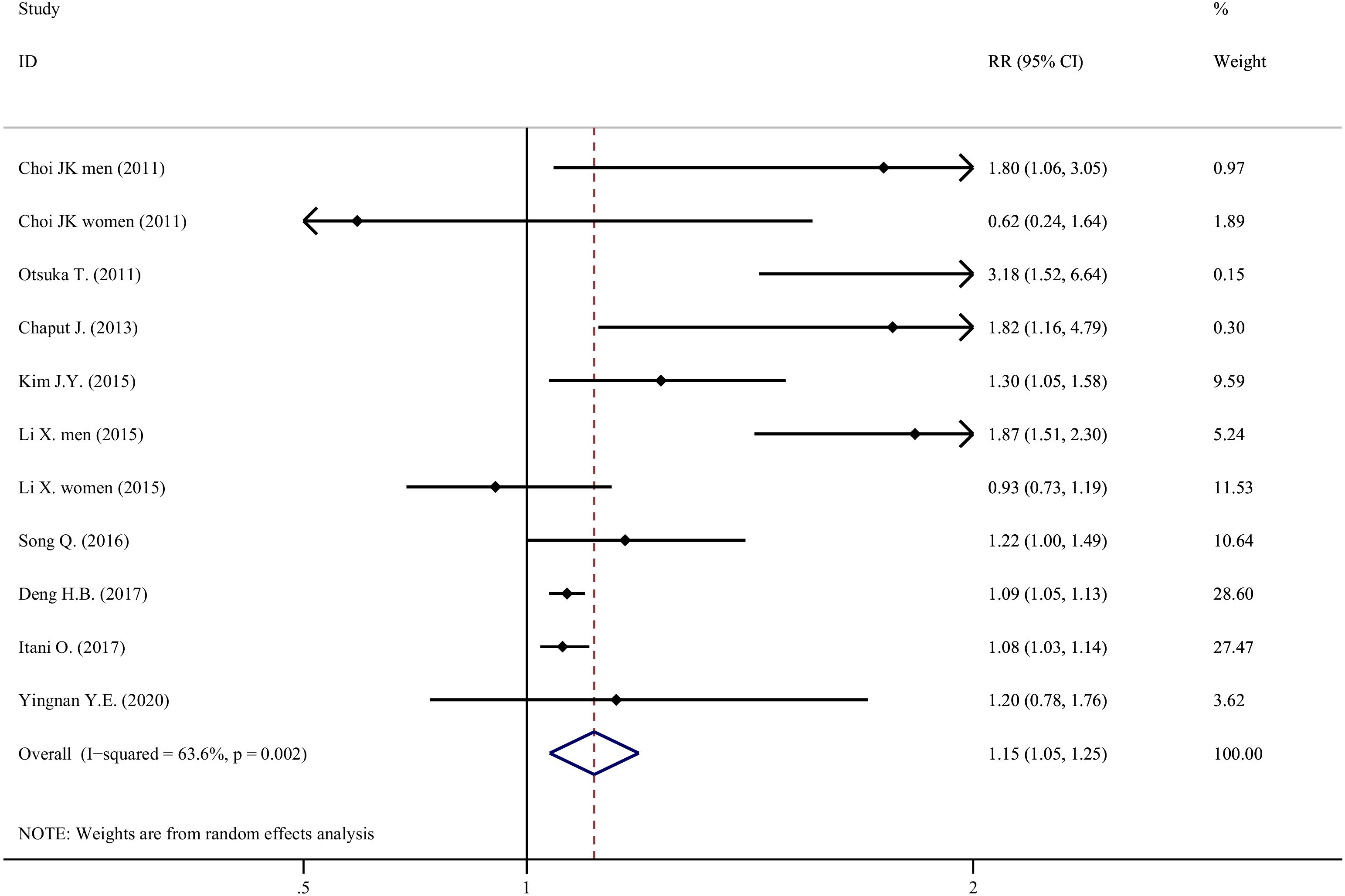
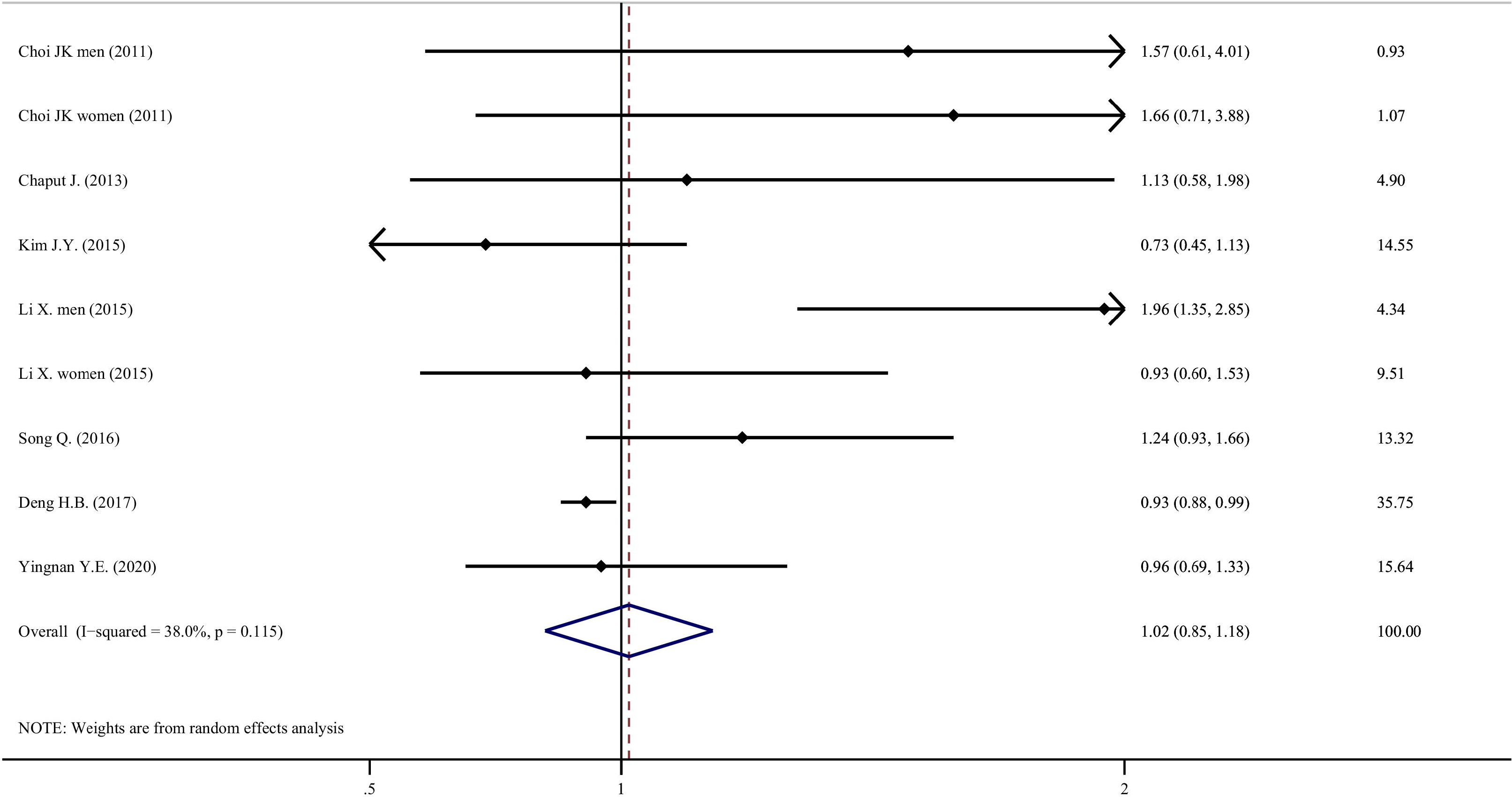
Forest plot of association between sleep duration and the risk of metabolic syndrome. (A) Comparing short sleepers with normal sleepers; (B) Comparing long sleepers with normal sleepers.

Compared with moderate sleep duration, the association between long sleep duration and risk of metabolic syndrome was not statistically significant, with an RR of 1.02 (95% CI=0.85-1.18, P=0.491, I^2^=38.0%, N=9; Figure 2B), using random effect model. The RR reduced to 0.94 (95% CI=0.89-0.99, P=0.050, I^2^=38.0%, N=9), using fixed effect model.

Among the seven studies that examined the effect of long sleep duration, six did not observe a significant association. Only Li X. found that long sleep duration increased the risk of metabolic syndrome among men (adjusted HR=1.96, 95% CI=1.35-2.85).

#### 3.3.2 Sleep duration and the prevalence of metabolic syndrome

Compared with moderate sleepers, people with short or long sleep duration had a higher prevalence of metabolic syndrome. The pooled OR of metabolic syndrome in short sleepers compared to moderate sleepers was 1.06 (95% CI=1.01-1.11, P<0.001, I^2^=66.5%, N=32; Figure 3A). The pooled OR of metabolic syndrome in long sleepers compared to moderate sleepers was 1.11 (95% CI=1.04-1.17, P<0.001, I^2^=73.8%, N=31; Figure 3B).

**Figure 3.**
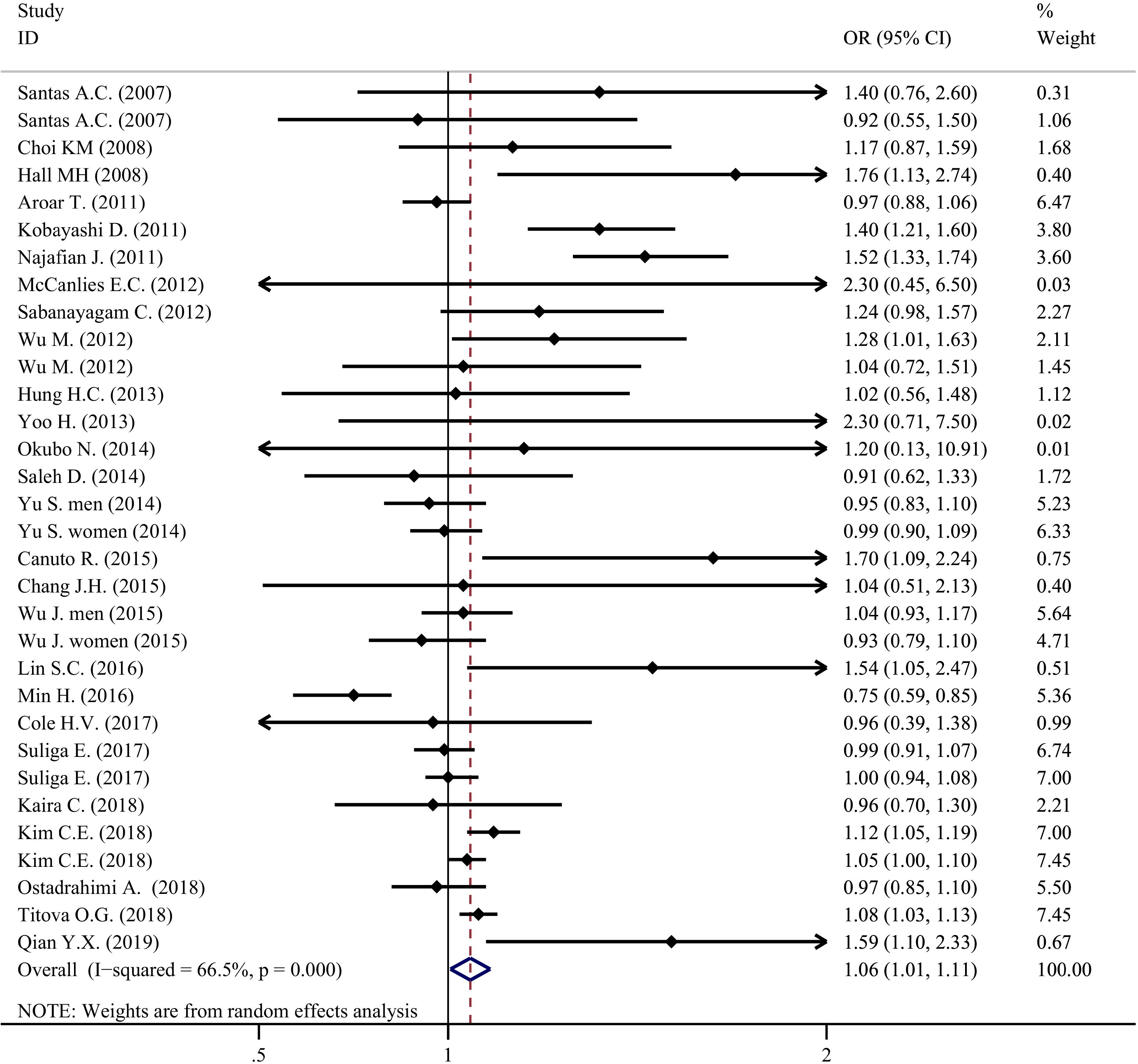
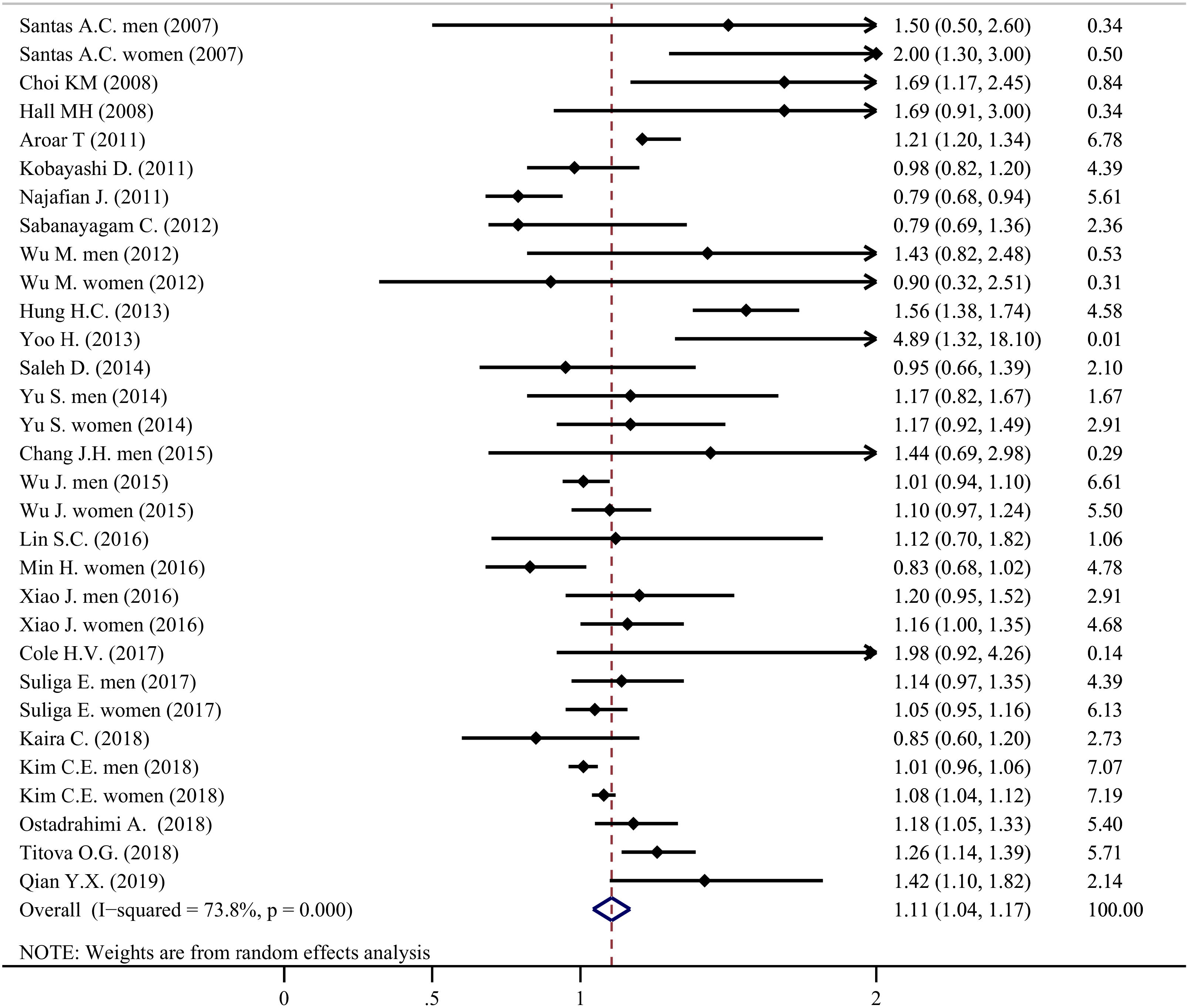
Forest plot of association between sleep duration and the prevalence of metabolic syndrome. (A) Comparing short sleepers with normal sleepers; (B) Comparing long sleepers with normal sleepers.

### 3.4 Possible publication bias for primary analysis

Results of Begg and Mazumdar and Egger test were shown in Table S4. No significant publication bias was observed. “Trim and fill” test indicated that the primary results was still significant after the missing studies were filled in (Table S5). Visual inspection of the funnel plots also did not reveal apparent publication bias (Figures not shown).

### 3.5 Subgroup analysis

Subgroup analysis of the cross-sectional studies were shown in Table 2. No significant subgroup difference was found for gender, continent, and definition of abnormal sleep duration. There were statistically significant subgroup effects (P<0.05 for heterogeneity between groups). For short sleepers, the specific domains were study population, sleep measurement, measures of metabolic syndrome and sample size. For long sleepers, the specific domains were sleep measurement, long sleep duration, measures of metabolic syndrome, sample size, and study quality. However, there was still unexplained heterogeneity within some subgroups. Therefore, the subgroup couldn’t fully explain the overall heterogeneity.

**Table 2.**
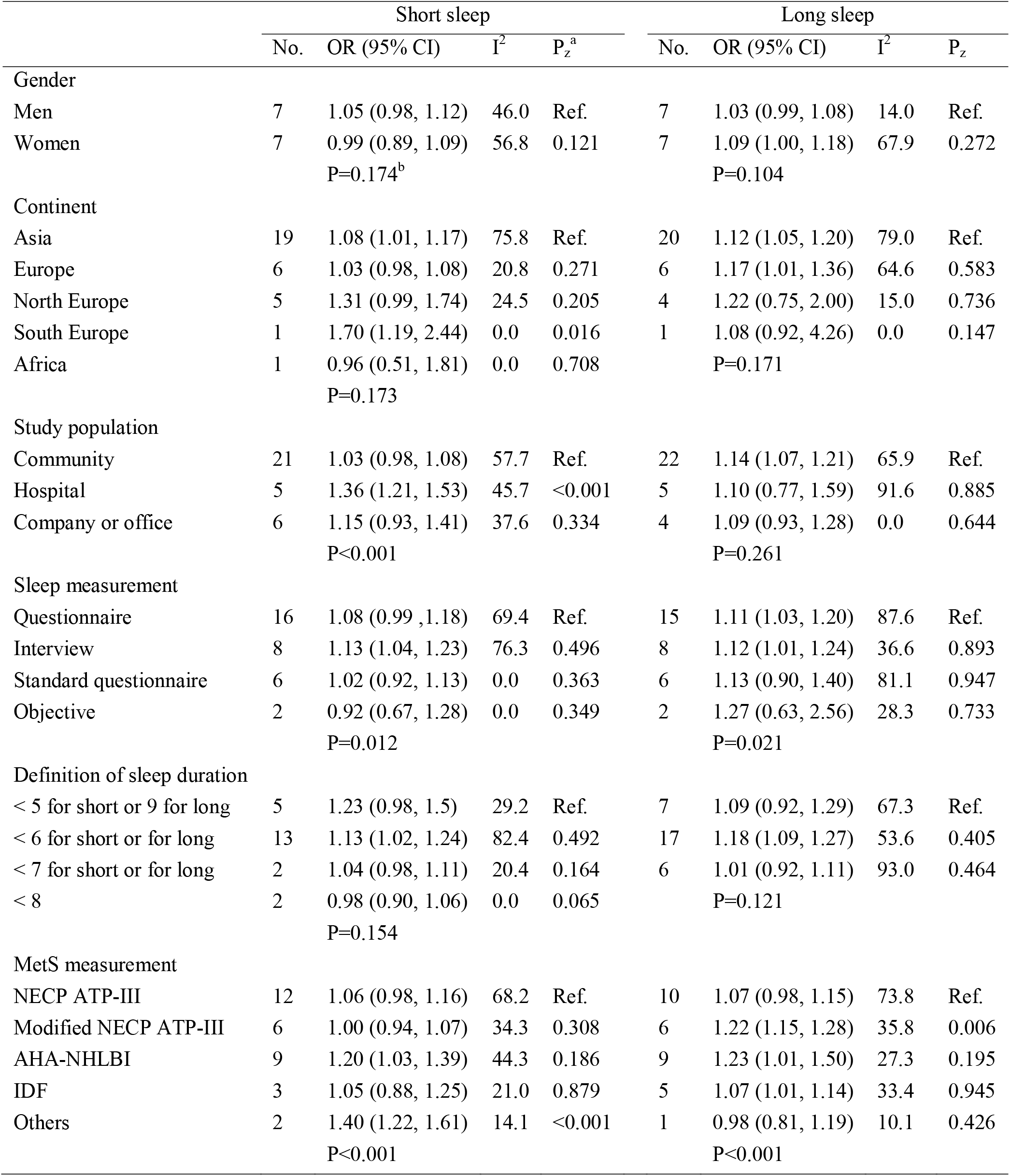
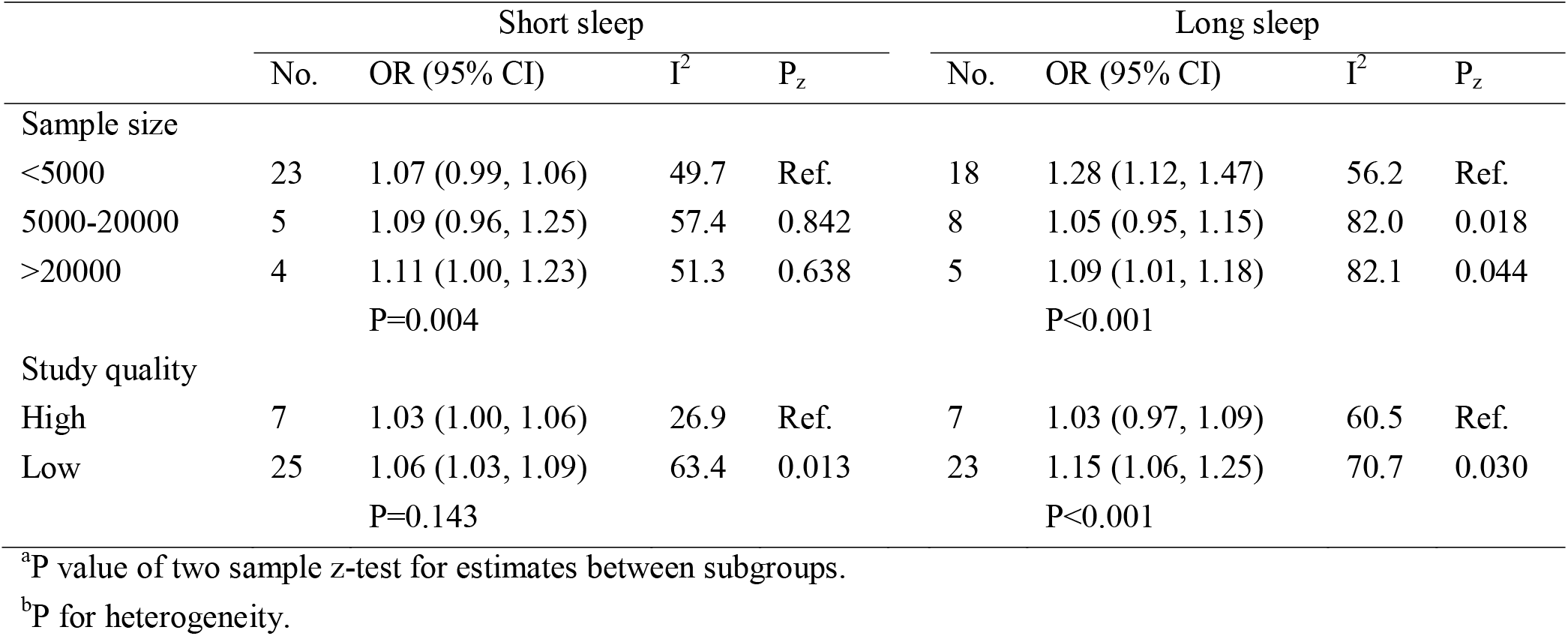
Subgroup meta-analysis of cross-sectional studies.

|  | Short sleep |  |  |  | Long sleep |  |  |  |
| --- | --- | --- | --- | --- | --- | --- | --- | --- |
|  | No. | OR (95% CI) | I<sup>2</sup> | P<sub>z</sub><sup>a</sup> | No. | OR (95% CI) | I<sup>2</sup> | P<sub>z</sub> |
| Gender |  |  |  |  |  |  |  |  |
| Men | 7 | 1.05 (0.98, 1.12) | 46.0 | Ref. | 7 | 1.03 (0.99, 1.08) | 14.0 | Ref. |
| Women | 7 | 0.99 (0.89, 1.09) | 56.8 | 0.121 | 7 | 1.09 (1.00, 1.18) | 67.9 | 0.272 |
|  |  | P=0.174<sup>b</sup> |  |  |  | P=0.104 |  |  |
| Continent |  |  |  |  |  |  |  |  |
| Asia | 19 | 1.08 (1.01, 1.17) | 75.8 | Ref. | 20 | 1.12 (1.05, 1.20) | 79.0 | Ref. |
| Europe | 6 | 1.03 (0.98, 1.08) | 20.8 | 0.271 | 6 | 1.17 (1.01, 1.36) | 64.6 | 0.583 |
| North Europe | 5 | 1.31 (0.99, 1.74) | 24.5 | 0.205 | 4 | 1.22 (0.75, 2.00) | 15.0 | 0.736 |
| South Europe | 1 | 1.70 (1.19, 2.44) | 0.0 | 0.016 | 1 | 1.08 (0.92, 4.26) | 0.0 | 0.147 |
| Africa | 1 | 0.96 (0.51, 1.81) | 0.0 | 0.708 |  | P=0.171 |  |  |
|  |  | P=0.173 |  |  |  |  |  |  |
| Study population |  |  |  |  |  |  |  |  |
| Community | 21 | 1.03 (0.98, 1.08) | 57.7 | Ref. | 22 | 1.14 (1.07, 1.21) | 65.9 | Ref. |
| Hospital | 5 | 1.36 (1.21, 1.53) | 45.7 | <0.001 | 5 | 1.10 (0.77, 1.59) | 91.6 | 0.885 |
| Company or office | 6 | 1.15 (0.93, 1.41) | 37.6 | 0.334 | 4 | 1.09 (0.93, 1.28) | 0.0 | 0.644 |
|  |  | P<0.001 |  |  |  | P=0.261 |  |  |
| Sleep measurement |  |  |  |  |  |  |  |  |
| Questionnaire | 16 | 1.08 (0.99, 1.18) | 69.4 | Ref. | 15 | 1.11 (1.03, 1.20) | 87.6 | Ref. |
| Interview | 8 | 1.13 (1.04, 1.23) | 76.3 | 0.496 | 8 | 1.12 (1.01, 1.24) | 36.6 | 0.893 |
| Standard questionnaire | 6 | 1.02 (0.92, 1.13) | 0.0 | 0.363 | 6 | 1.13 (0.90, 1.40) | 81.1 | 0.947 |
| Objective | 2 | 0.92 (0.67, 1.28) | 0.0 | 0.349 | 2 | 1.27 (0.63, 2.56) | 28.3 | 0.733 |
|  |  | P=0.012 |  |  |  | P=0.021 |  |  |
| Definition of sleep duration |  |  |  |  |  |  |  |  |
| < 5 for short or 9 for long | 5 | 1.23 (0.98, 1.5) | 29.2 | Ref. | 7 | 1.09 (0.92, 1.29) | 67.3 | Ref. |
| < 6 for short or for long | 13 | 1.13 (1.02, 1.24) | 82.4 | 0.492 | 17 | 1.18 (1.09, 1.27) | 53.6 | 0.405 |
| < 7 for short or for long | 2 | 1.04 (0.98, 1.11) | 20.4 | 0.164 | 6 | 1.01 (0.92, 1.11) | 93.0 | 0.464 |
| < 8 | 2 | 0.98 (0.90, 1.06) | 0.0 | 0.065 |  | P=0.121 |  |  |
|  |  | P=0.154 |  |  |  |  |  |  |
| MetS measurement |  |  |  |  |  |  |  |  |
| NECP ATP-III | 12 | 1.06 (0.98, 1.16) | 68.2 | Ref. | 10 | 1.07 (0.98, 1.15) | 73.8 | Ref. |
| Modified NECP ATP-III | 6 | 1.00 (0.94, 1.07) | 34.3 | 0.308 | 6 | 1.22 (1.15, 1.28) | 35.8 | 0.006 |
| AHA-NHLBI | 9 | 1.20 (1.03, 1.39) | 44.3 | 0.186 | 9 | 1.23 (1.01, 1.50) | 27.3 | 0.195 |
| IDF | 3 | 1.05 (0.88, 1.25) | 21.0 | 0.879 | 5 | 1.07 (1.01, 1.14) | 33.4 | 0.945 |
| Others | 2 | 1.40 (1.22, 1.61) | 14.1 | <0.001 | 1 | 0.98 (0.81, 1.19) | 10.1 | 0.426 |
|  |  | P<0.001 |  |  |  | P<0.001 |  |  |
|  | No. | OR (95% CI) | I<sup>2</sup> | P<sub>z</sub> | No. | OR (95% CI) | I<sup>2</sup> | P<sub>z</sub> |
| Sample size |  |  |  |  |  |  |  |  |
| <5000 | 23 | 1.07 (0.99, 1.06) | 49.7 | Ref. | 18 | 1.28 (1.12, 1.47) | 56.2 | Ref. |
| 5000-20000 | 5 | 1.09 (0.96, 1.25) | 57.4 | 0.842 | 8 | 1.05 (0.95, 1.15) | 82.0 | 0.018 |
| >20000 | 4 | 1.11 (1.00, 1.23) | 51.3 | 0.638 | 5 | 1.09 (1.01, 1.18) | 82.1 | 0.044 |
|  |  | P=0.004 |  |  |  | P<0.001 |  |  |
| Study quality |  |  |  |  |  |  |  |  |
| High | 7 | 1.03 (1.00, 1.06) | 26.9 | Ref. | 7 | 1.03 (0.97, 1.09) | 60.5 | Ref. |
| Low | 25 | 1.06 (1.03, 1.09) | 63.4 | 0.013 | 23 | 1.15 (1.06, 1.25) | 70.7 | 0.030 |
|  |  | P=0.143 |  |  |  | P<0.001 |  |  |

<sup>a</sup>P value of two sample z-test for estimates between subgroups.
<sup>b</sup>P for heterogeneity.

For short sleepers, hospital-based participants had a higher OR than the community-based. With regard to definition of short sleep duration, there was a tendency that lower duration was associated with higher OR. Nevertheless, the P value was not significant.

For long sleepers, compared with “NECP” group, “modified-NECP” group had higher OR. Studies with more participants or higher quality had lower OR.

### 3.6 Sensitivity analysis

None of the sensitivity analysis substantially altered the effect of both long and short sleep duration on metabolic syndrome (Figure S1).

### 3.7 Meta-regression

With regard to cross-sectional studies, a multivariate meta-regression analysis (Table 3) was conducted to examine the potential influence of different factors on the natural logarithm of the OR of short or long sleep duration with prevalence of MetS. For short sleepers, shorter definition of duration was associated with higher OR (P=0.011 for multivariable test, and P=0.099 for univariable test). Higher study quality was associated with lower OR (P=0.010 for univariable test and P=0.033 for multivariable test). The effect of mean age was significant. However, the clinical effect (coef=-0.01) was little. For long sleepers, none of the study factors was significant.

**Table 3.** Results of meta-regression analysis of cross-sectional studies.

| Short sleep duration |  |  |  |  |
| --- | --- | --- | --- | --- |
|  | Univariable analysis |  | Multivariable analysis |  |
|  | Coef. | P value | Coef. | P value |
| Mean age, years | -0.01 (-0.02, -0.01) | 0.020 | -0.01 (-0.01, 0.00) | 0.086 |
| Proportion of males, % | 0.08 (-0.10, 0.27) | 0.356 | -0.05 (-0.23, 0.14) | 0.622 |
| Definition of short/long sleep | -0.07 (-0.15, 0.01) | 0.099 | -0.08 (-0.15, -0.12) | 0.011 |
| Sample size | 0.01 (-0.08, 0.09) | 0.864 | 0.01 (-0.09, 0.09) | 0.351 |
| Study quality | -0.15 (-0.30, 0.00) | 0.039 | -0.26 (-0.61, 0.09) | 0.149 |
| Long sleep duration |  |  |  |  |
|  | Univariable analysis |  | Multivariable analysis |  |
|  | Coef. | P value | Coef. | P value |
| Mean age, years | 0.00 (0.00, 0.01) | 0.113 | 0.00 (-0.00, 0.01) | 0.720 |
| Proportion of males, % | 0.01 (-0.18, 0.02) | 0.891 | -0.04 (-0.44, 0.35) | 0.828 |
| Definition of short/long sleep | -0.16 (-0.33, 0.10) | 0.776 | -0.09 (-0.28, 0.10) | 0.958 |
| Sample size | -0.09 (-0.18, 0.00) | 0.056 | -0.08 (-0.25, 0.08) | 0.318 |
| Study quality | -0.01 (-0.14, 0.10) | 0.807 | 0.01 (-0.50, 0.52) | 0.360 |

## 4 Discussion

To our knowledge, this was the first meta-analysis on the association between sleep duration and the risk of metabolic syndrome. Our study indicated that short sleep duration correlated with an increase in the risk of metabolic syndrome. Long duration was not statistically associated with the risk of metabolic syndrome. With regard to cross-sectional studies, both short and long sleep duration was associated with higher prevalence of metabolic syndrome.

Our results differed from that of three previous meta-analysis, which reported the association between sleep duration and metabolic syndrome. Limited by sample size, the three studies did not distinguish cross-sectional studies and cohort studies in the main outcome. *Ju 2013* and *Iftikhar 2015* reported only short sleep duration was associated with metabolic syndrome^7,8^, while *Xi 2014* linked both short and long sleep duration to metabolic syndrome^9^. Moreover, we observed narrower confidence intervals, which might be due to our higher number of included datasets. Nowadays, a growing number of articles reported an inverted-U shaped association between sleep duration and health outcomes, including metabolic syndrome^22^. Our findings suggested that long sleep duration might have no effect.

Compared with the previous studies, we further conducted comprehensive sub-group analysis and meta-regression for cross-sectional studies. In subgroup analysis, using z test, we found no statistically significant differences in subgroups of sex, continent, and sleep measurement. For short sleepers, hospital-based participants had a higher prevalence than community-based participants, which was consistent with a previous meta-analysis. One possible reason was that people in hospital had poor health conditions. The results of subgroup analysis and meta-regression showed that studies with high quality or larger sample size had smaller risk estimates. In multivariable meta-regression, shorter sleep duration was linearly associated with higher prevalence of metabolic syndrome. Longer sleep duration had no linear association. This “J-shaped” association was not consistent with the “U-shaped” association between sleep duration reported in many articles^22^. However, we thought this was not contradictory. In the meta-regression, “sleep duration” was a cut-off point defined by different authors from different ethnicities; and in one specific study, the author calculated the association among participants from a same ethnic.

Several mechanisms linked sleep duration to metabolic syndrome. Short sleep duration could lead to the following endocrine changes, by affecting carbohydrate metabolism, hypothalamo-pituitary-adrenal axis and sympathetic activity. Decreased glucose tolerance and insulin sensitivity would raise the glucose levels; increased levels of ghrelin, decreased levels of leptin and increased appetite correlated with higher waist circumstance; increased cortisol concentrations were associated with higher blood pressure^23,24^. Short sleepers tended to have low-grade inflammation. Elevated levels of high-sensitivity C-reactive protein and IL-6 correlated with cardiovascular events^25^. Long sleep duration was linked to sleep fragmentation, which would cause plenty of health outcomes, including metabolic change^26^. Long sleepers also have less time to take excise, which might contribute to the association^27^. Both short and long sleep duration had bidirectional association with circadian rhythm, which is a risk factor for metabolic disorders^28,29^. Despite these mechanisms which could explain the association, whether sleep duration was a causal risk factor for metabolic syndrome was still not sure^30^. Cohort studies still could not provide a causal result, though they have more power than cross-sectional studies. To prove a causal relationship, we need further examine the effect of changes in sleep duration^31^, and product Mendelian randomization study, a method using measured variation in genes.

Our meta-analysis summarized the currently evidence of the association between abnormal sleep duration and the risk of metabolic syndrome, and indicated that short sleep duration was a risk factor for new onset metabolic syndrome. Strength of our study were the comprehensive search and strict inclusion criteria. However, there were some limitations that should be taken into consideration. First, the sleep categories varied between studies. There was no unified standard for definition of abnormal sleep duration. Second, the estimates of relative risk had less accuracy than that in the individual patient data meta-analysis. Third, we only included nine cohort studies, which prevent us from conducting further research, such as subgroup analysis and meta-regression. Fourth, only two studies used objective sleep measurement. More studies based on the objective measurement are needed in the future.

## 5 Conclusions

Both short and long sleep duration was associated with high prevalence of metabolic syndrome cross-sectionally. Short sleep duration, rather than long sleep duration, was associated with a significant increase in risk for metabolic syndrome. Sufficient sleep duration should be recommended to prevent metabolic syndrome.

## Data Availability

This is a meta-analysis

## 6 Author contributions

Qi Fang and Jianian Hua contributed to the conception and design of the study. Jianian Hua organized the database. Jianian Hua and Hezi Jiang performed the statistical analysis. Jianian Hua wrote the first draft of the manuscript. Qi Fang and Hezi Jiang reviewed the manuscript. All authors approved the final version of the paper.

## 9 Preprint

